# Estimating life expectancy and years of life lost in epidemiological studies: a review of methods using an example from multimorbidity

**DOI:** 10.1101/2022.01.18.22269472

**Authors:** Yogini V Chudasama, Kamlesh Khunti, Clare L Gillies, Nafeesa N Dhalwani, Melanie J Davies, Thomas Yates, Francesco Zaccardi

## Abstract

**Background and objective:** There has been an increasing interest in using life expectancy metrics, such as years of life lost (YLL), to explore epidemiological associations. YLL is easier to understand for both healthcare professionals and the lay people and has become a common measure for evaluating public health priorities. As the literature presents a range of approaches to estimate it, this review aims to: (1) summarise the key methods; (2) show how to implement them using current software; (3) apply them in a real-world example.

**Methods:** We investigated simpler nonparametric as well as parametric, model-based methods to estimate of YLL, including: (1) Years of potential life lost (YPLL); (2) Global Burden of Disease (GBD) approach; (3) Chiang’s life tables; (4) Epi-demographic approach; and (5) Flexible Royston-Parmar parametric survival model. We used data from the UK Biobank with baseline measures collected in 2006-2010 and linkage to mortality records. We selected 36 chronic conditions: participants with two or more conditions were categorised as having multimorbidity.

**Results:** For the YPLL and GBD method, the analytical procedures allow only to quantify the average YLL within each group (with and without multimorbidity) and, from them, their difference. Conversely, for the Chiang’s life tables, the epi-demographic approach, and the Royston-Parmar survival model, both the remaining life expectancy within each group and the YLL could be estimated. In 499,992 UK Biobank participants (white ethnicity, 94%; women, 55%) with a median (IQR) age of 58 (50-63) years, 98,605 (20%) had multimorbidity and 11,871 deaths occurred during the follow-up. The YLLs comparing subjects with vs without multimorbidity varied significantly according to the technique and the modelling approach used: from a longer life expectancy in subjects with multimorbidity using the YPLL and the GBD method to a shorter one using the other three methods (i.e., at 65 years, the YLL were 1.8, 1.3, and 4.6 years using Chiang’s, epi-demographic, and Royston-Parmar approach, respectively).

**Conclusions:** When comparing the burden of a disease on life expectancy across studies caution is needed as methods may estimate different quantities. While deciding among different methods to estimate YLL, researchers should consider such differences in relation to the purpose of the research and the type of available data.

SUMMARY BOX

- The concept of years of life lost (YLL) is easier to understand compared to traditional estimates from survival analysis, such as hazard ratios, but very few studies report it.
- A range of different methods of estimating YLL are reviewed: from basic methods – such as life tables – to most recent and advanced methods using statistical modelling.
- Using the example of multimorbidity, the estimated numbers of YLL differs between methods, as each method focused on estimating different quantities.
- This review will help promote a better understanding and use of life expectancy and YLL metrics in a wide range of studies in health care research.

## INTRODUCTION

Recently, there has been an increasing interest in using life expectancy measures, such as years of life lost (YLL), as metrics to explore epidemiological associations. Life expectancy estimates are indeed easier to understand and can be simply translated to deliver public health priorities and messages.^1, 2^ Yet, the majority of epidemiological studies using time-to-event analysis still present results using the most common metric of hazard ratios (HRs).

This is a relative measure, with the potential advantage of being more transportable, i.e. can be applied to populations different from those where they have been estimated,^3, 4^ and requires fewer assumptions compared to some methods for estimating YLL. However, HRs can be difficult to understand and frequently misinterpreted;^5^ furthermore, large (or small) HRs may translate into negligible absolute differences.^3^

A better understanding of the impact of an exposure can be achieved by considering also absolute differences. The term “YLL” broadly encompasses a range of different definitions, names, methods, and quantities, including loss in expectation of life, loss of life expectancy, life lost, lost-lifetime, excess years of life lost, and difference in remaining or residual life expectancy.^6–13^ Differences among these metrics depend on a number of factors, such as: (i) the reference group for the comparisons with those having the condition of interest (either the standard life expectancy from the general population – assuming they are free from the condition – or those without the condition in the same (observed) population); (ii) the starting age (from birth, commonly used in demography; or from a specific age – at disease diagnosis or at death among those with the disease); ^2^ (iii) the population used for the estimates (based only on those individuals who have died, or based on extrapolation beyond the last observed follow-up as most studies do not follow all the patients to the end of life).^10^

One approach to estimate YLL is based on quantifying the mean survival time, obtained by constructing the survival curve: the area under the curve, with respect to age, corresponds to the mean age at death; for two populations – one including individuals with a certain characteristic or condition (i.e., cancer, diabetes, multimorbidity) and the other including subjects without such condition – two survival curves can be obtained: the difference between the areas of the two survival curves indicates the modelled difference in the age at death associated with the presence of the condition (i.e., YLL).

To our knowledge, there has not been an overall review of YLL methods most commonly used in epidemiological studies, their relative advantages and disadvantages, and the software available to compute relevant metrics. In this context, our study aims to summarise and illustrate the differences and interpretations between the most common methods of estimating YLL and apply them to a clinically-relevant example of multimorbidity (i.e. the presence of two or more chronic conditions) using the UK Biobank, a large contemporary cohort; examples of different methods are complemented by statistical codes.

### YEARS OF LIFE LOST: DEFINITIONS and METHODS

From reviewing the literature, we identified the most common methods to estimate life expectancy/YLL in epidemiological investigations, from simpler nonparametric to more advanced parametric, model-based methods, reflecting in part the progressively larger access to individual-level rather than aggregate data. We give a brief summary of each method before comparing and contrasting them using multimorbidity as an example.

Although these methods allow to quantify the difference in the years of life in relation to the presence of a condition, we note that the underlying estimands are different, thus we do not *a-priori* expect consistent findings across all the explored methods. Further details about the methods, the analytical procedures, and the statistical codes are reported in the **Supplementary Material.**

## 1. Years of potential life lost

Years of potential life lost (YPLL) is calculated as the difference between the age at death and age from a selected cut-off, e.g. 65, 75, or 85 years. This concept dates back to the 1940s – when it was first introduced to compare deaths due to tuberculosis with heart disease and cancer – and, for each death, the number of years of life remaining up until premature mortality was calculated.^14^ For example, if premature mortality is taken to be at 65 years, an individual who has died at 55 years has lost 10 years of potential life. Deaths after the cut-off age are ignored, giving more value and weight to deaths occurring at younger ages.^6^ The data for this method is restricted to only those who have died. A disadvantage of using YPLL is that the selected cut-off age is arbitrary, making it difficult to compare results across studies. However, there have been further adaptations of standardising YPLL to overcome this issue.^6^ Another disadvantage is that accurate information on the cause of death is required.^10^ YPLL is usually based on ranking data by the leading causes of death for a given period of time: as such, the results emphasise the causes of death most commonly found in the younger population.^6^

### Software

YPLL is simple to compute. This method can be applied using a calculator or an Excel spreadsheet.

## 2. Global Burden of Disease approach

The Global Burden of Disease (GBD) approach is an extension of the YPLL measure and, although restricted to those who have died, this method includes deaths beyond an arbitrary cut-off. It is based on comparing the age of death to an external standard life expectancy curve, and can further incorporate time discounting and age weighting.^7, 15^ Since the YPLL method can be overly sensitive to deaths at younger ages, the GBD approach uses therefore a discounting strategy based on the idea that the productive years of life are more valuable than the very young or the very old. From the introduction of the GBD study in 1993,^16^ the burden of disease concept has expanded to numerous countries and is used by major health organisations, such as the World Health Organization (WHO), as well as an outcome measure in many cost-effectiveness analysis.^16^ The general equation for the GBD YLL approach is developed on key assumptions of standardised weightings for age, discount rate, and constants. For each age group, it requires details on the number of participants, number of deaths, and the standard life expectancy. The benefit of using the GBD approach is that the age-adjusted rates allow comparisons of causes of deaths in rank order across groups, conditions, and over time,^15^ which is important from a public health perspective because it identifies patterns of preventable YLL.

### Software

An open source programming code is available in R that allows to estimate GBD YLL:^7^ all that is required is a value of the number of deaths, average age of death, and the standard life expectancy for that age.^17^ Alternatively, an Excel template created by the WHO National tools allows the calculation to be carried out for all ages at the same time.^18^

## 3. Chiang’s life tables

Life tables provide a description of the mortality rates and are one of the most important tools used in demography and actuarial science. The two forms of life tables are: cohort or period/current life tables. The cohort life table identifies the actual observed mortality experience of individuals born at the same time followed up throughout time, and is similar to national life table data. The period/current life table describes the mortality experience of a population subject to the age-specific mortality rates currently observed and applied to a hypothetical cohort, and any future changes to mortality rates would not be taken into account, which is used in epidemiological studies. There are a number methods to construct life tables;^19–21^ the most common is the Chiang’s life table method,^8^ as this approach is widely used for population estimates (i.e., by the Office for National Statistics (ONS)) in the UK^17, 21^ and has been applied to large epidemiological studies.^22^ Chiang’s life table method usually groups the data by five year age intervals, yet this depend on the size of the study.

The life table is then constructed with the number of participants and deaths in each age interval, and is based on conditional probability of surviving to the start of the age group. The survival probabilities are applied to a hypothetical cohort and using cumulative distribution the expected number of years is estimated. The population can be stratified into groups where two life tables are constructed separately and the expected remaining years of life is subtracted to find YLL. The results from this method have been found to be less stable in smaller populations, and the chosen value of the ending age interval is arbitrary, therefore making it difficult to produce reliable estimates and compare results between studies.^19, 20^ Nevertheless, life tables are one of the oldest and more traditional methods of calculating remaining life expectancy, and works well in large national studies and applied to YLL analyses.

### Software

Templates are freely available online from the ONS,^17^ or the Association of Public Health Epidemiologists in Ontario.^23^ They can also be calculated using R,^24^ SAS,^25^ or Stata.^26^

## 4. Epi-demographic approach: Epi package

By linking a demographic and survival analysis approach within a parametric modelling framework, YLL can be estimated using the Epi package in R.^27^ Firstly, the follow-up time is split in small intervals: this results in multiple observations for the same subject, with age, time into the study, and calendar time increasing by the same amount: for example, a subject starts the study/observation at age 45.5 years in year 2000.5 with time into study split in intervals of six months (0.5 years): thus, at the end of the next interval, age will be 46 years, calendar time 2001, and the time into the study 0.5 years.^28^ Age is then modelled with a Poisson regression (with a natural spline) using each small time interval into the study as offset: the estimated coefficients allow the prediction of age-specific rates and, from them, the survival functions/curves; the area under the curve allows to estimate the residual life. This method has been used for large national registries,^9, 29^ and it is straightforward to use.

### Software

The Epi package has the functions to calculate estimated residual life (*erl*) and the YLL *(yll*); it also allows modelling the possible transition from a state without to a state with the disease/condition of interest across which the YLL is computed.^30^ This method has also been used in Stata.^31^

## 5. Flexible Royston-Parmar parametric survival model

The Royston-Parmar flexible parametric survival model allows for greater flexibility to accurately capture data and extrapolate for future predictions.^32^ This is done by using restricted cubic splines thus obtaining smooth parametric survival curve.^32^ It was first introduced by Royston and Parmar in 2002 as an alternative to the Cox proportional model and implemented in Stata.^32, 33^ The most recent user-written Stata command *stpm2* is a quick and efficient method to fit flexible parametric models.^34^ Previous studies have used this method mainly in cancer epidemiology.^35–37^ There are a number of possible outputs available from these models: this review focuses on calculating YLL. Firstly, *stpm2* is used to fit the regression model using age as time scale and including the condition of interest as covariate.^38^ Then, the postestimation *predictnl* command enables the estimation of the survival curves across age; calculating the area under the curve is then straightforward, and YLL is calculated as the area between the two survival curves of interest. The advantages of using this method is that it performs better for smaller samples compared to some of the previously reported methods;^39^ it enables 95% confidence intervals estimation (with delta method) around YLL; and potential confounders can be accounted for (both conditional and average/standardised estimation).^40^ The disadvantage of this method is the computationally intensive process, particularly for large datasets and when confidence intervals are required.

### Software

This method is very broad and there are a number of functions that can be applied.^31^ As well as having commands available in Stata, it can also be carried out in SAS^41^ and R.^42^ In this review, we will consider its implementation in Stata.

### EXAMPLE FROM MULTIMORBIDITY

This real-world example focuses on estimating the number of YLL in a population of individuals with multimorbidity compared to those without multimorbidity. This study is reported in line with the Strengthening the Reporting of Observational Studies in Epidemiology (STROBE) guideline (**Supplementary Checklist S1**).

### Data

We used UK Biobank data (Application Number 14146), one of the world’s largest Biobank cohorts, designed to improve the prevention, diagnosis, and treatment of chronic diseases in middle-aged adults, recruited from 22 sites across England, Wales and Scotland. This longitudinal study included 502,629 participants with baseline measures collected between 2006 and 2010 and linkage to mortality records.^43^ Written informed consent was obtained prior to data collection and ethics was approved by the North-West Research Ethics Committee.^44^ Overall, 91 participants withdrew from the study. The deaths occurring within the first two years at baseline were removed (n=2,516) to reduce the possible impact of reverse causation, i.e. participants with undiagnosed or sub-clinical disease(s) leading to mortality. There was a small number of participants before the age of 45 years at follow-up (n=30, of which 29 died) who were excluded from analyses to avoid uncertainty of deaths at younger age (**Supplementary Figure S1**).

### Mortality

All-cause mortality data were obtained from the National Health Service (NHS) Information Centre for participants from England and Wales and the NHS Central Register for participants from Scotland. Time for survivors was censored on 31^st^ January 2016 for England and Wales and 30^th^ November 2015 for Scotland.

### Definition of multimorbidity

The UK Biobank collected self-reported medical information based on physician diagnosis at the baseline assessment. Three sources of selecting long-term cardiovascular, non-cardiovascular, or mental health conditions was used. The first included conditions from the Quality and Outcomes Framework which reports the most common diseases in the UK;^45^ the second from a large UK based study, containing 40 of the recommended core disorders for any multimorbidity measure;^46^ and the last from a systematic review on multimorbidity indices that included 17 conditions.^47^ Based on these sources, we selected a total of 36 chronic conditions. Participants with two or more of these 36 chronic conditions were categorised as having multimorbidity (**Supplementary Methods S1**).

### Statistical methods

Baseline characteristics were stratified by multimorbidity status and summarised. To model the association between multimorbidity and all-cause mortality, unadjusted Cox proportional hazards regression was used with age as time scale. HRs and corresponding 95% confidence intervals (95% CI) were calculated. Participants without multimorbidity constituted the reference category. For each YLL method described above, we provide detailed steps and example statistical code in the **Supplementary Material Methods S2-S6** to demonstrate their implementation. Stata version 17.0, R studio version 1.4, and Microsoft Excel were used to perform the analyses.

## RESULTS

### Baseline characteristics

A total of 499,992 participants were included in this study, with most participants being white (94%) and women (55%); the median (IQR) age was 58 (50-63) years. Subjects with multimorbidity (n=98,605; 19.7%) were older compared to participants without (>60 years: 52% vs. 35%, respectively; **Table 1**).

**Table 1.**
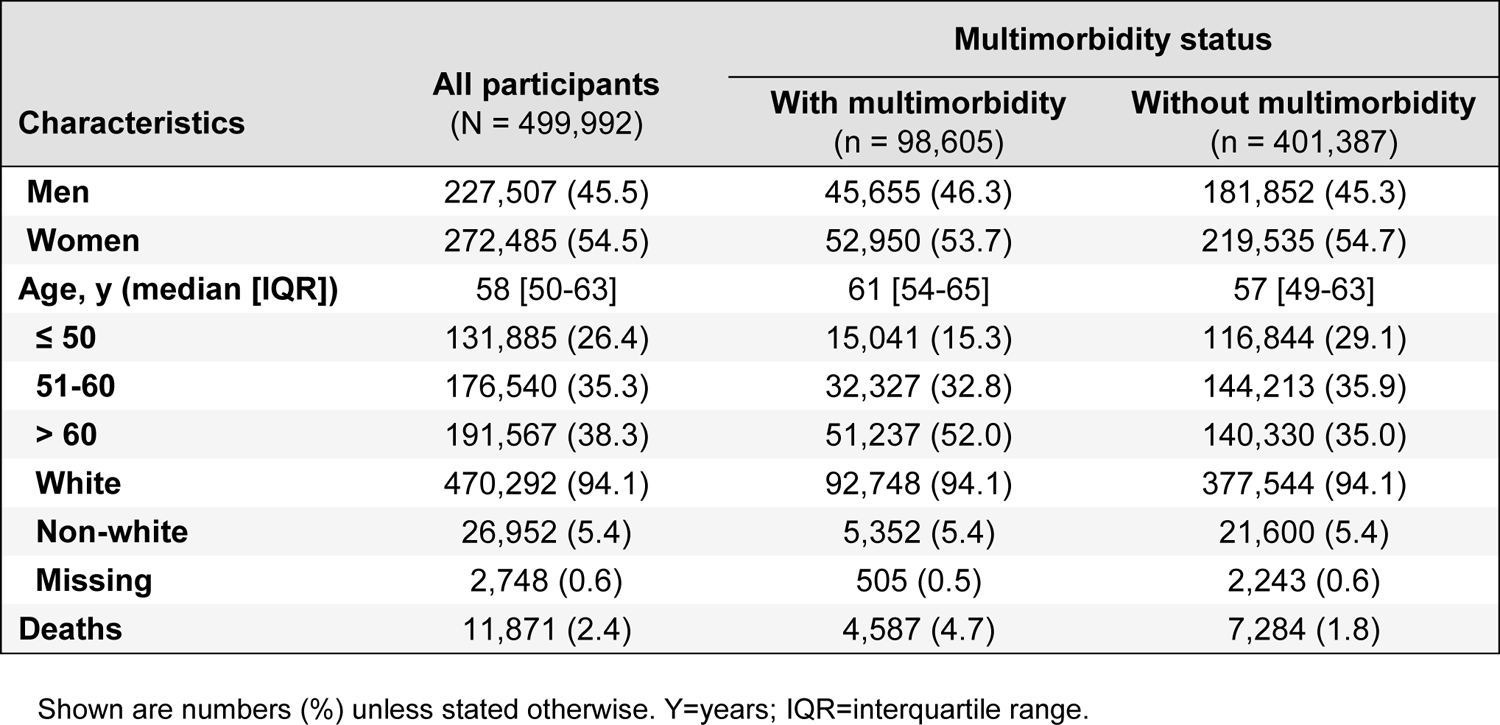
Participant characteristics and outcomes by multimorbidity status

| Characteristics | All participants<br>(N = 499,992) | Multimorbidity status |  |
| --- | --- | --- | --- |
|  |  | With multimorbidity<br>(n = 98,605) | Without multimorbidity<br>(n = 401,387) |
| **Men** | 227,507 (45.5) | 45,655 (46.3) | 181,852 (45.3) |
| **Women** | 272,485 (54.5) | 52,950 (53.7) | 219,535 (54.7) |
| **Age, y (median [IQR])** | 58 [50-63] | 61 [54-65] | 57 [49-63] |
| **≤ 50** | 131,885 (26.4) | 15,041 (15.3) | 116,844 (29.1) |
| **51-60** | 176,540 (35.3) | 32,327 (32.8) | 144,213 (35.9) |
| **> 60** | 191,567 (38.3) | 51,237 (52.0) | 140,330 (35.0) |
| **White** | 470,292 (94.1) | 92,748 (94.1) | 377,544 (94.1) |
| **Non-white** | 26,952 (5.4) | 5,352 (5.4) | 21,600 (5.4) |
| **Missing** | 2,748 (0.6) | 505 (0.5) | 2,243 (0.6) |
| **Deaths** | 11,871 (2.4) | 4,587 (4.7) | 7,284 (1.8) |

Shown are numbers (%) unless stated otherwise. Y=years; IQR=interquartile range.

### Multimorbidity and all-cause mortality

During a mean study follow-up of 7 (range 2-10) years and 3.5 million person-years at risk, 11,871 deaths were recorded: 4,587 (4.7%) in subjects with multimorbidity and 7,284 (1.8%) in those without. The median [IQR] age at death was 61 [54-65] and 57 [49-63] years in subjects with and without multimorbidity, respectively (**Supplementary Figure S2**). Using age as the time scale, the Cox regression model showed that the mortality rates were twice as high for those with multimorbidity compared to those without, with an unadjusted hazard ratio of 2.00 (95% CI: 1.93, 2.08).

### Years of life lost estimates across methods

The results for the five methods are presented in **Table 2**. Given the analytical procedures underpinning the estimations, for the YPLL and GBD method only the average YLL could be estimated within each group (multimorbidity and no multimorbidity): then, from them, the difference between the two groups could be estimated. Conversely, for the Chiang’s life tables, the epi-demographic approach with the *Epi* package, and the flexible Royston-Parmar parametric survival approach with *stpm2,* both the remaining life expectancy within each group and the YLL could be estimated (**Figure 1**).

**Table 2.** Estimating years of life lost in subjects with multimorbidity compared to those without multimorbidity using different methods

| Method | Age (years) | Average years of life lost (SD) |  | Difference in average years of life lost, without vs with |
| --- | --- | --- | --- | --- |
|  |  | With multimorbidity | Without multimorbidity |  |
| Years of potential life lost | 65 cut-off | 6.15 (4.81) | 6.88 (4.95) | 0.73 |
|  | 75 cut-off | 8.52 (6.05) | 9.67 (6.64) | 1.15 |
| Global burden of disease approach | 45-49 | 21.61 | 21.63 | 0.01 |
|  | 65-69 | 13.80 | 13.85 | 0.05 |
| Method | Age (years) | Remaining life expectancy (95% CI) |  | Years of life lost (95% CI), without vs with |
|  |  | With multimorbidity | Without multimorbidity |  |
| Chiang's life tables | 45-49 | 19.99 (19.30, 20.67) | 27.54 (27.36, 27.73) | 7.56 |
|  | 65-69 | 9.72 (9.60, 9.84) | 11.55 (11.49, 11.61) | 1.83 |
| Epi-demographic approach | 45 | 46.25 | 51.16 | 4.91 |
|  | 65 | 31.54 | 32.82 | 1.27 |
| Royston-Parmar PH | 45 | 39.52 (38.85, 40.19) | 44.71 (44.03, 45.39) | 5.19 (4.81, 5.58) |
|  | 65 | 21.05 (20.38, 21.71) | 25.63 (24.95, 28.89) | 4.59 (4.24, 4.93) |
| Royston-Parmar non PH | 45 | 40.34 (39.35, 41.33) | 43.97 (43.05, 44.88) | 3.63 (2.27, 4.99) |
|  | 65 | 22.19 (21.21, 23.17) | 24.82 (23.89, 28.56) | 2.62 (1.27, 3.98) |

Years of life lost = without multimorbidity – with multimorbidity.
SD=standard deviation; CI=confidence interval; PH=proportional hazards

**Figure 1.**
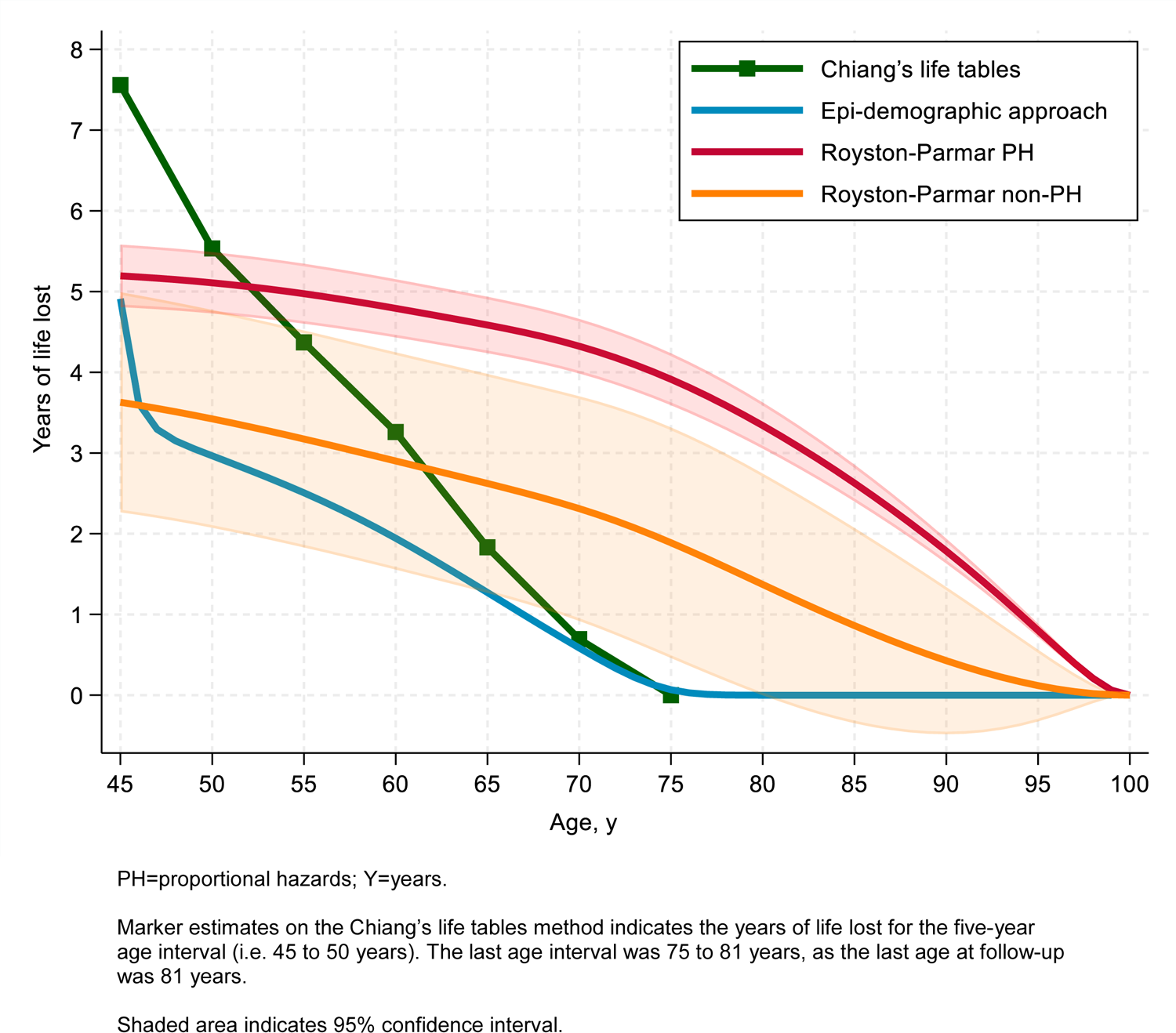
Number of years of life lost in subjects with multimorbidity compared to those without in the UK Biobank

## 1. Years of potential life lost (Methods S2)

Using a selected cut-off age of 65 years, there was a total of 25,504 potential years of life lost. The average YPLL lost for a subject with multimorbidity was 6.15 (SD, 4.81) years and for a subject without 6.88 (SD 4.95), giving a difference of 0.73 years. Using a selected cut-off point of 75 years, corresponding figures were: 103,552, 8.52 (SD, 6.05), 9.67 (SD, 6.64), and 1.15 years, respectively. Since there was a higher number of deaths in subjects without vs with multimorbidity (61.4% vs. 38.6%), the results showed larger YPLL values in those without multimorbidity. In addition, the baseline age for the majority of those with multimorbidity was over 60 years (52%) compared to those without multimorbidity (35%); therefore, when followed-up until the age of death, participants with multimorbidity were less likely to lose more years of potential life than those without multimorbidity.

## 2. Global Burden of Disease approach (Methods S3)

The GBD approach estimate of the total YLL was 62,538 and 103,595 years in subjects with and without multimorbidity, respectively. For participants who died between 45-49 years, the average YLL was 21.61 years in those with multimorbidity and 21.63 years in those without: the difference in YLL was therefore 0.01 years. For participants who died between 65-69 years, corresponding figures were 13.80, 13.85, and 0.05 years.

## 3. Chiang’s life tables approach (Methods S4)

Using Chiang’s life tables, the results showed that, at the age of 45-49 years, the remaining life expectancy was 19.99 (95% CI: 19.30, 20.67) years in participants with multimorbidity and 27.54 (27.36, 27.73) in those without: the YLL was therefore 7.56 years. At the age of 65-69 years, corresponding figures were: 9.72 (9.60, 9.84), 11.55 (11.49, 11.61), and 1.83 years.

## 4. Epi-demographic approach: Epi package (Methods S5)

The results using the *Epi* package showed that, at the age of 45 years, the remaining life expectancy in subjects with multimorbidity was 46.25 years and in those without 51.16 years, resulting in YLL of 4.91 years. At the age of 65 years, corresponding estimates were 31.54, 32.82, and 1.27 years.

## 5. Flexible Royston-Parmar parametric survival model (Methods S6)

The flexible Royston-Parmar parametric survival model showed that, at the age of 45 years, participants with multimorbidity had 39.52 (95% CI: 38.85, 40.19) remaining years of life while those without 44.71 (44.03, 45.39) years, resulting in YLL of 5.19 (4.81, 5.58) years.

Corresponding figures at the age of 65 years were: 21.05 (20.38, 21.71), 25.63 (24.95, 28.89), and 4.59 (4.24, 4.93) years. When the effect of multimorbidity was assumed not constant over time (i.e., non-proportionality of hazards), the results were: at 45 years, 40.34 (39.35, 41.33) remaining years of life in subjects with multimorbidity and 43.97 (95% CI: 43.05, 44.88) those without, resulting in YLL of 3.63 (2.27, 4.99) years; at 65 years, 22.19 (21.21, 23.17) in and 24.82 (23.89, 28.56) subjects with and without multimorbidity, respectively, resulting in YLL of 2.62 (1.27, 3.98) years.

## DISCUSSION

The YLL measures are becoming a key health indicator in understanding a population’s health. In this article, a range of different methods for estimating YLL were reviewed, from the oldest – such as YPLL and life tables – to the most recent and advanced method using statistical modelling. The example demonstrated that the total numbers of YLL due to multimorbidity varied according to the technique used, as each method focuses on estimating different quantities, emphasising the need to understand the different concepts and use the most suitable method for the database and the specific research question. In turn, our results also underline the need to consider such differences when comparing (and summarising) studies reporting “years of life lost” related to a certain condition.

An overview of the YLL methods described in our manuscript is illustrated in **Figure 2**. The simplest method is the YPLL, which is especially useful in a clinical setting requiring minimal data input. Though, YPLL is usually found in large studies that help quantify the total disease burden and identify the most common causes of premature deaths. When applied to our multimorbidity example, the results showed a gain in YLL, since the baseline age for the majority of those with multimorbidity was much older than those without multimorbidity; therefore, participants with multimorbidity were less likely to lose more YPLL than those without multimorbidity at age of death. The GDB approach, which additionally accounted for time discounting and age weighting, showed a small difference in the average YLL. These two methods are based on only participants who have died and do not consider people who are alive during the time period; it is thus most useful, probably, in extremely large national databases.

**Figure 2.**
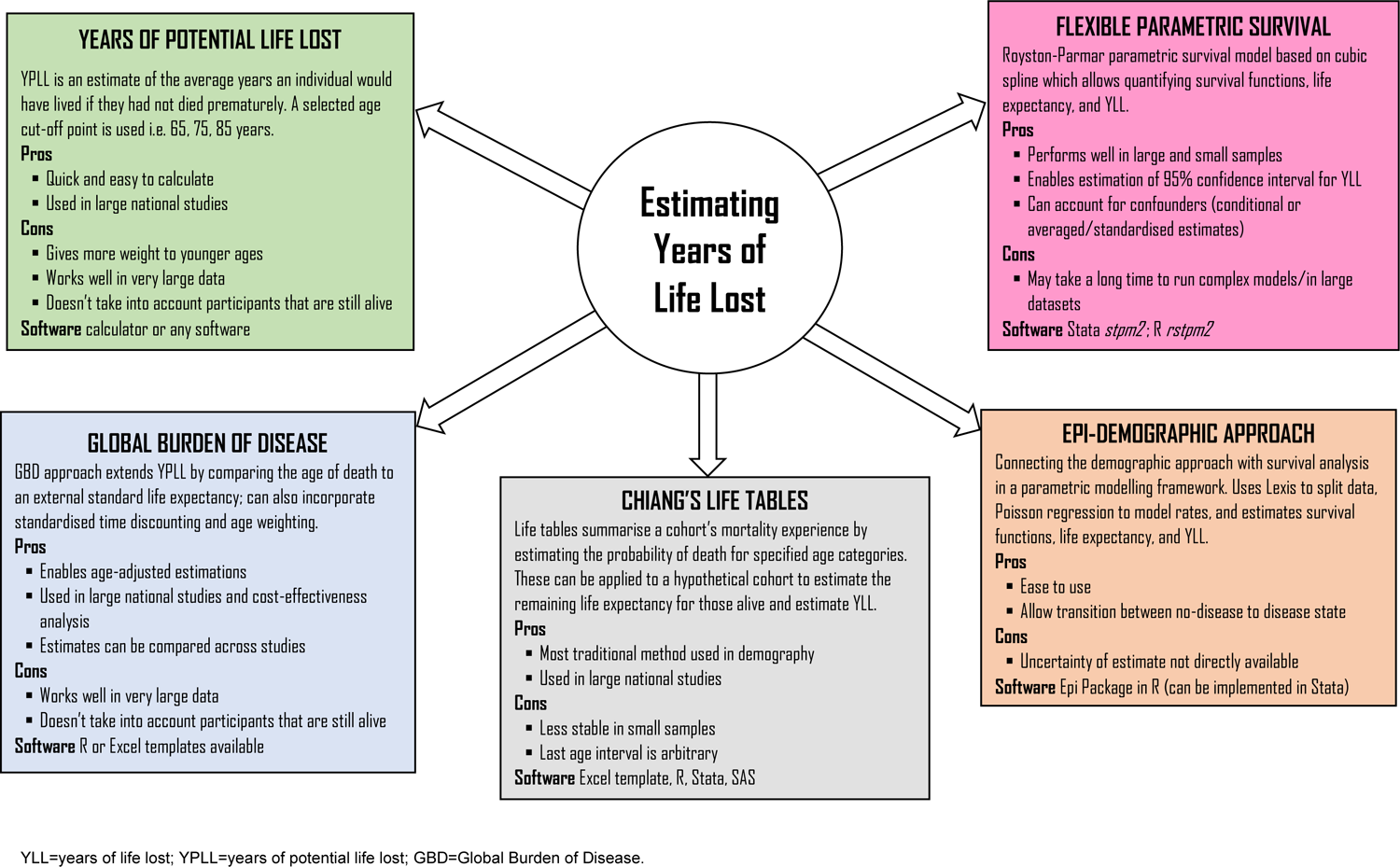
Summary of methods to estimate years of life lost

The results from Chiang’s life tables showed a higher number of YLL only at the youngest age – at 45-49 years, YLL was 7.56 years, compared to the epi-demographic approach (at 45 years, the YLL was 4.91 years) or the Royston-Parmar model (at 45 years, the YLL was 5.19 years) – and then decreases. This is because: (1) the results were based on aggregated data where abridged life tables with five year age intervals were used, as the sample size was not large enough for individual year data (unabridged or complete life table),^20^ and the mortality rate in each age interval is assumed to be constant; (2) the chosen value for the last age interval was 7, which was the difference between 75 (inclusive) and the last observed age, 81 years. This value is arbitrary and varies in the literature, as any age interval value could be applied, or it could be left open-ended.^19, 20, 48^ Leaving this open-ended interval assumes a constant hazard of death for the remaining ages, which may lead to erroneous conclusions about mortality measures.^20^ Therefore, in relation to the type of life table (abridged or unabridged) and the last age interval value, the results for the remaining life expectancies and YLL from the Chiang’s life table method will vary.

The epi-demographic approach using the *Epi* package is based on survival curves obtained from mortality rates, following a Poisson modelling on split data: this fully parametric approach gives more flexibility (i.e., may allow multiple time scales and/or other covariates for conditional estimates). The results from the example showed higher remaining life expectancies than other methods, as this was extrapolated to 100 years; of note, the smaller the extrapolation, the smaller the remaining life expectancy (i.e., the area under the curve). Additionally, the estimated survival curve depends also on the functional form of age in the Poisson regression (i.e., continuous vs natural cubic spline – with possible effects in relation to the number and the position of the knots). An advantage of the *Epi* package is the possibility to easily model the transition from one group to the other exposure group across which YLL is estimated, in our case from no multimorbidity to multimorbidity.

The final method is based on the Royston-Parmar model implemented in the *stpm2* command in Stata, which enables to estimate the survival functions (or differences among them) and their uncertainties. Similar to the *Epi* package, the results of this fully parametric approach may reflect decisions around the maximum age used for the survival prediction (in this analysis we used 100 years as for the Poisson modelling approach); the number/position of the knots for the cubic spline; or interactions between the covariate of interest and age. Besides conditional estimates, i.e. for specific values of the covariates included in the model, recent further development also enables marginal (standardised) average effect estimates, including YLL.^40^ This method also allowed the estimate of the 95% CI, giving a measure of uncertainty. Notably, running the full code was time consuming.

Modelling approaches have advantages in that they allow for greater flexibility for both individual-level and grouped data.^49^ As most studies do not follow participants to the end of life, the risk of extrapolating (i.e. to 100 years) needs to be taken into consideration. This means that factors such as the time-scale, length of follow-up, number of participants and deaths, as well as the age distribution of the population and among those who died, can influence the results. To date, there is no standard method for estimating life expectancy and YLL, and the most appropriate method may vary depending on the research question being addressed and the available data.

From the example presented here, the mortality rates were twice as high for those with multimorbidity compared to those without (HR, 2). Using the Royston-Parmar model, the YLL at the age of 45 years in participants with multimorbidity was on average 5.2 years lower than in participants without; one advantage of presenting HRs is that, as a relative measure, the estimate is more transportable and can be used to compare and combine estimates from different populations with different baseline risk.^3, 4^ In this perspective, a recent study confirmed that the HR estimates from the UK Biobank are similar to those obtained from studies based on the general population.^50^ On the other hand, presenting the absolute risk in terms of ‘years of your life’ may be far more influential from a public heath perspective and policy makers, as the concept is easier to grasp.^1^ Therefore, presenting both relative and absolute measures strengthens the reporting of associations from an epidemiological, clinical, and public perspective, and aligns with current guidelines for conducting and reporting epidemiological studies (STROBE; item #16).^51^

This review has some limitations. We focused only on the most commonly used YLL methods; others methods (or further extension of those mentioned in this study) are available and may also enable the estimation of the YLL by different causes of death.^52–54^ UK Biobank participants were volunteers, with slightly higher representation from affluent groups; therefore, participants may not be completely representative of the UK population.^55^ Participants who died within the first two years of follow-up were excluded to reduce the risk of reverse causation, though it is still possible that participants with multimorbidity may generally be less well, which could result in a higher mortality rate. Additionally, data on chronic diseases was collected at baseline, yet participants may have subsequently developed other chronic conditions throughout follow-up. As most studies predict survival beyond the duration of the study data, further investigations with longer follow-up may explore the impact of the model structural uncertainty on the YLL estimates.^56^ Lastly, the minimum sample size of participants and the number of events required to estimate life expectancy and YLL for each method is still yet to be clarified, and further research is needed, possibly through simulation studies.^20^

Strengths of this study include the utilisation and illustration of different methods to estimate life expectancy and YLL, with statistical codes reported for the available software. Furthermore, we applied the methods to an emerging clinical and research area, as one of the four recommendations for research from the National Institute for Health and Care Excellence (NICE) guidance for multimorbidity focuses on predicting the life expectancy.^57^ Finally, the methodologies used to estimate the number of YLL could be also be used to enhance the interpretation and potentially the promotion of beneficial interventions, such as gain in disease-free life expectancy,^58^ or lifestyle modifications.^59, 60^ For instance, patients with multimorbidity may be motivated to exercise more if their doctors communicated a simple health care message, such as 10 minutes of brisk walking a day could lead you to having up to 4 years of additional life.^60^

### RECOMMENDATIONS

YLL metrics are simple summary measures that can enhance the interpretation of the findings from epidemiological studies for health care professionals and the public. When possible, we therefore recommend that, alongside a relative risk (i.e. rate or hazard ratio), also the absolute risk (i.e. number of events, rates, YLL) should be reported, thus providing a more complete and actionable information. As the available methods to estimate YLL metrics could relate to different quantities, comparing the spatiotemporal burden of diseases on the life expectancy may be difficult. Furthermore, investigators should not only choose the YLL method in view of the purpose of the research, the type of available data, and the required flexibility but – when a modelling strategy is considered – they should also assess the consistency of the findings across different models.

## DECLARATIONS

### Author contributions

KK, FZ and ND conceived the idea of the study; ND acquired the data; YC carried out the review and statistical analysis; FZ supervised the analysis. YC, FZ, KK, and CG interpreted the findings; and YC drafted the manuscript. YC had full access to all the data in the study and takes responsibility for the integrity of the data and the accuracy of the data analysis. All authors critically reviewed the manuscript and YC revised the manuscript for final submission.

## LIST OF ABBREVIATIONS

CI: Confidence intervals

GBD: Global Burden of Disease

HRs: Hazard ratios

ONS: Office for National Statistics

QoF: Quality and outcomes framework

STROBE: Strengthening the Reporting of Observational Studies in Epidemiology

UK: United Kingdom

WHO: World Health Organization

YLL: Years of life lost

YPLL: Years of potential life lost

## Data Availability

The data that support the findings of this study are available from UK Biobank project site, subject to registration and application process. Further details can be found at https://www.ukbiobank.ac.uk.

## Acknowledgements

This research has been conducted using the UK Biobank Resource (Reference 14614). We would like to acknowledge Dr Sarwar Islam Mozumder for his comments on an initial draft of the manuscript. We acknowledge the support from the National Institute for Health Research (NIHR) Applied Research Collaboration East Midlands (ARC EM), and the NIHR Leicester Biomedical Research Centre. The views expressed are those of the author(s) and not necessarily those of the NIHR or the Department of Health and Social Care.

## Ethical approval and informed consent

All participants gave written informed consent prior data collection. UK Biobank has full ethical approval from the NHS National Research Ethics Service (16/NW/0274). UK Biobank Reference14614.

## Supplementary material

**Figure S1.**
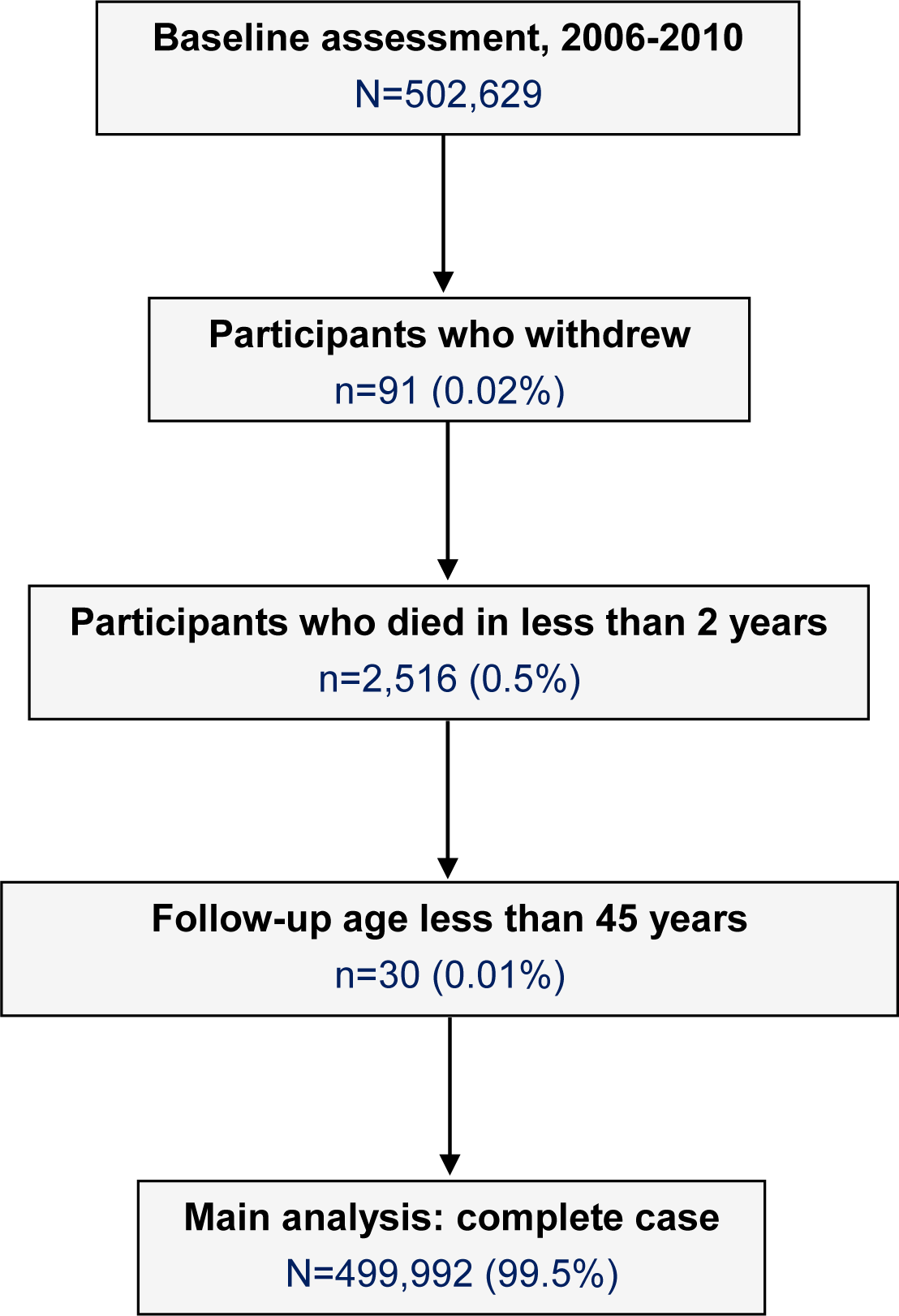
Flow chart of participants included in the study

**Figure S2.**
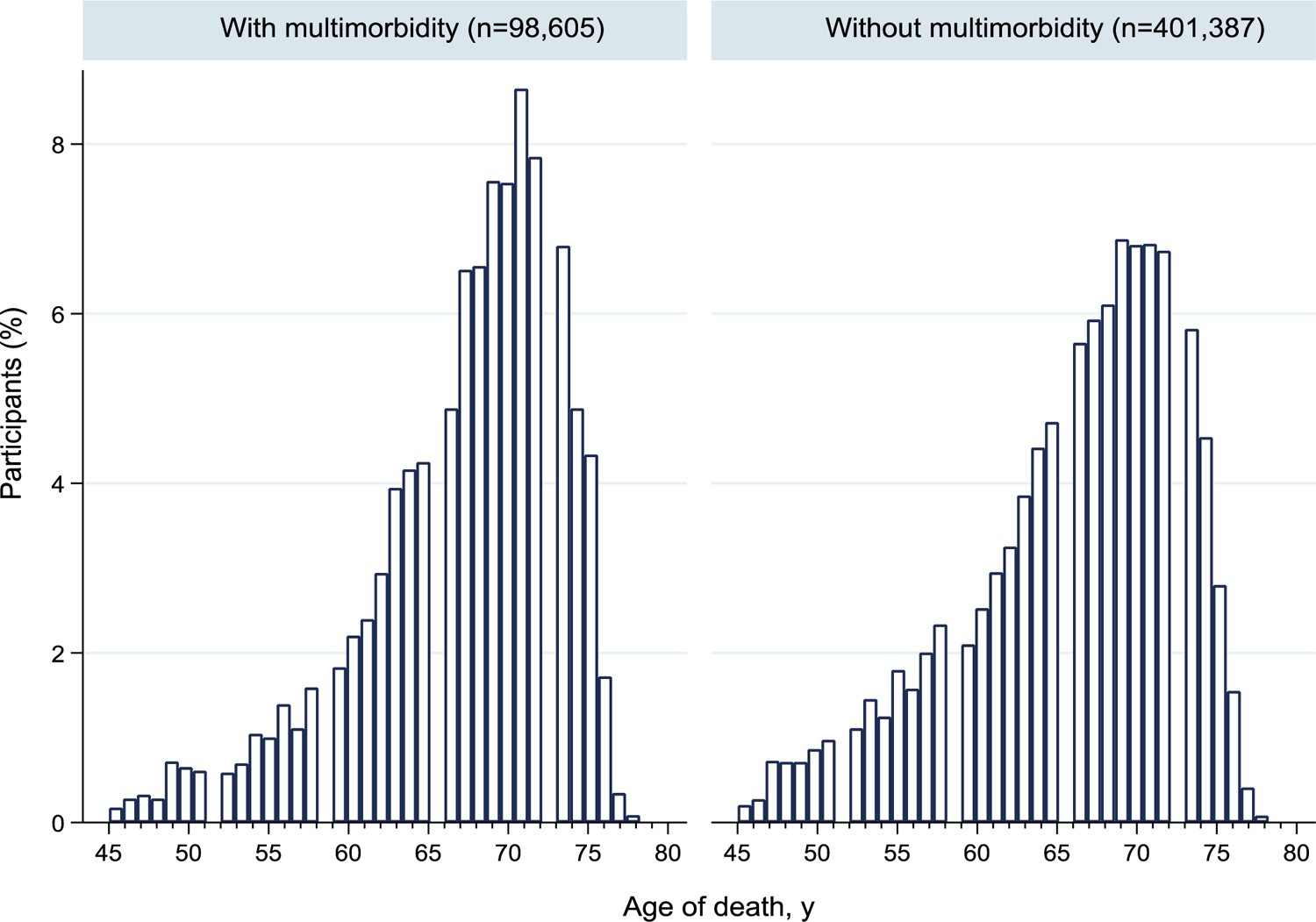
Distribution of age of death by multimorbidity status

## Methods S1. List of the 36 chronic conditions included in the definition of multimorbidity

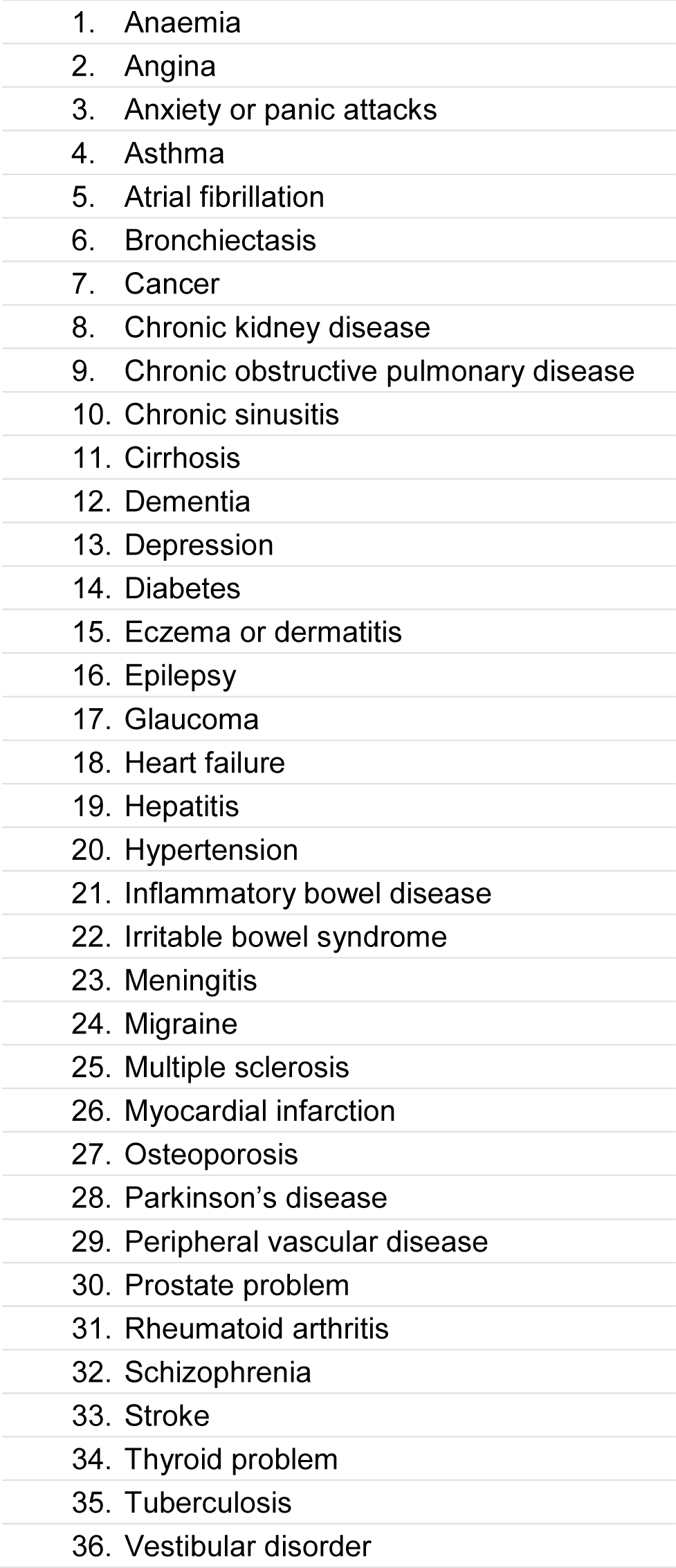

|  |
| --- |
| 1. Anaemia |
| 2. Angina |
| 3. Anxiety or panic attacks |
| 4. Asthma |
| 5. Atrial fibrillation |
| 6. Bronchiectasis |
| 7. Cancer |
| 8. Chronic kidney disease |
| 9. Chronic obstructive pulmonary disease |
| 10. Chronic sinusitis |
| 11. Cirrhosis |
| 12. Dementia |
| 13. Depression |
| 14. Diabetes |
| 15. Eczema or dermatitis |
| 16. Epilepsy |
| 17. Glaucoma |
| 18. Heart failure |
| 19. Hepatitis |
| 20. Hypertension |
| 21. Inflammatory bowel disease |
| 22. Irritable bowel syndrome |
| 23. Meningitis |
| 24. Migraine |
| 25. Multiple sclerosis |
| 26. Myocardial infarction |
| 27. Osteoporosis |
| 28. Parkinson's disease |
| 29. Peripheral vascular disease |
| 30. Prostate problem |
| 31. Rheumatoid arthritis |
| 32. Schizophrenia |
| 33. Stroke |
| 34. Thyroid problem |
| 35. Tuberculosis |
| 36. Vestibular disorder |

## Methods S2. Years of potential life lost example using UK Biobank

**Step 1.** Select an age cut-off point (i.e. 65, 75, 85 years).

**Step 2.** Exclude participants who have died or are still alive after the cut-off point.

**Step 3.** For each participant who died before the end point, calculate the YPLL by subtracting the age at death from the cut-off point.

YPLL _individual_ = cut-off point – age at death

*Example*

Selected cut-off = 65 Age at death = 55

YPLL individual = 65 - 55 = 10 years of potential life lost

**Step 4.** Total the individual YPLL.

**Steps 5.** To find the average YPLL, the total number of YPLL is divided by the total number of deaths.

**Steps 6.** The average YPLL for those with and without multimorbidity is subtracted to find the difference in YPLL.

*Example (10 subjects with multimorbidity and 8 without)*

| Multimorbidity |  |  |  | No Multimorbidity |  |  |  |
| --- | --- | --- | --- | --- | --- | --- | --- |
| Subject ID | Age at death | Cut-off | YPLL individual | Subject ID | Age at death | Cut-off | YPLL individual |
| 1 | 55 | 65 | 10 | 11 | 59 | 65 | 6 |
| 2 | 53 | 65 | 12 | 12 | 59 | 65 | 6 |
| 3 | 45 | 65 | 20 | 13 | 62 | 65 | 3 |
| 4 | 44 | 65 | 21 | 14 | 62 | 65 | 3 |
| 5 | 59 | 65 | 6 | 15 | 63 | 65 | 2 |
| 6 | 61 | 65 | 4 | 16 | 64 | 65 | 1 |
| 7 | 61 | 65 | 4 | 17 | 58 | 65 | 7 |
| 8 | 63 | 65 | 2 | 18 | 57 | 65 | 8 |
| 9 | 49 | 65 | 16 |  |  |  |  |
| 10 | 39 | 65 | 26 |  |  |  |  |
| Total |  |  | 121 | Total |  |  | 36 |

The example above shows an average of 121/10 = 12.1 YPLL in subjects with multimorbidity and 36/8 = 4.5 YPLL in those without; the multimorbidity vs no multimorbidity difference is therefore 12.1 - 4.5 = 7.6 YPLL.

## Methods S3. Global Burden of Disease approach example using UK Biobank

**Step 1.** Calculate the number of participants and deaths in each age interval (i.e., participants and deaths occurring at age 50, 51, 52……)

The number of deaths in each age interval was calculated for participants with and without multimorbidity, and by men and women. These numbers can be calculated using any software/calculator.

**Step 2.** Determine an external standard life expectancy, which is the average number of years that those aged *x* exact will live. This can be from the Coale and Demeny Model Life Tables West which is a set of standardised life expectancies, or from national registries such as the Office for National Statistics (ONS) in the UK.

In this study, the UK Biobank data was last censored on 31^st^ January 2016 for England and Wales and 30^th^ November 2015 for Scotland. We used the ONS National Life Tables by age and sex in 2013 to 2015 as the external standard life expectancy: this is based on population estimates from births and deaths by dates of registration for a period of 3 consecutive years. National life tables: UK https://www.ons.gov.uk/peoplepopulationandcommunity/birthsdeathsandmarriages/lifeexpectancies/datasets/nationallifetablesunitedkingdomreferencetables (Accessed 18-01-2022)

**Step 3.** Substitute the values into the general YLL formula:

The crude expected years of life lost is:

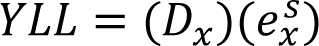

where *D_x_* is the number of deaths and *e^s^_x_* is the standard age of death from the external life expectancy.

We could simply use the crude expected YLL to estimate the number of YLL by each age and then find the total (following table).

*Example: female subjects with and without multimorbidity*

| With multimorbidity |  |  |  | Without multimorbidity |  |  |  |
| --- | --- | --- | --- | --- | --- | --- | --- |
| Age at death | No. of deaths ($D_x$) | Standard life expectancy ($e_x^s$) | YLL | Age at death | No. of deaths ($D_x$) | Standard life expectancy ($e_x^s$) | YLL |
| 50 | 17 | 34.2 | 581.40 | 50 | 8 | 34.20 | 273.60 |
| 51 | 19 | 33.28 | 632.32 | 51 | 10 | 33.28 | 332.80 |
| 52 | 18 | 32.35 | 582.30 | 52 | 12 | 32.35 | 388.20 |
| 53 | 15 | 31.43 | 471.45 | 53 | 18 | 31.43 | 565.74 |
| 54 | 34 | 30.52 | 1037.68 | 54 | 19 | 30.52 | 579.88 |
| Total | 103 |  | 3305.15 |  | 67 |  | 2140.22 |

The example above shows how to quantify the YLL for the age-group 50-54 years. The average of 3305.15/103 = 32.09 YLL in female subjects with multimorbidity and 2140.22/67 = 31.94 YLL in those without; the multimorbidity vs no multimorbidity difference is therefore 32.09 - 31.94 = 0.15 YLL.

To further extend the basic formula and incorporate the discounting and age weighting requires a range of assumptions.

The following general YLL formula is used:

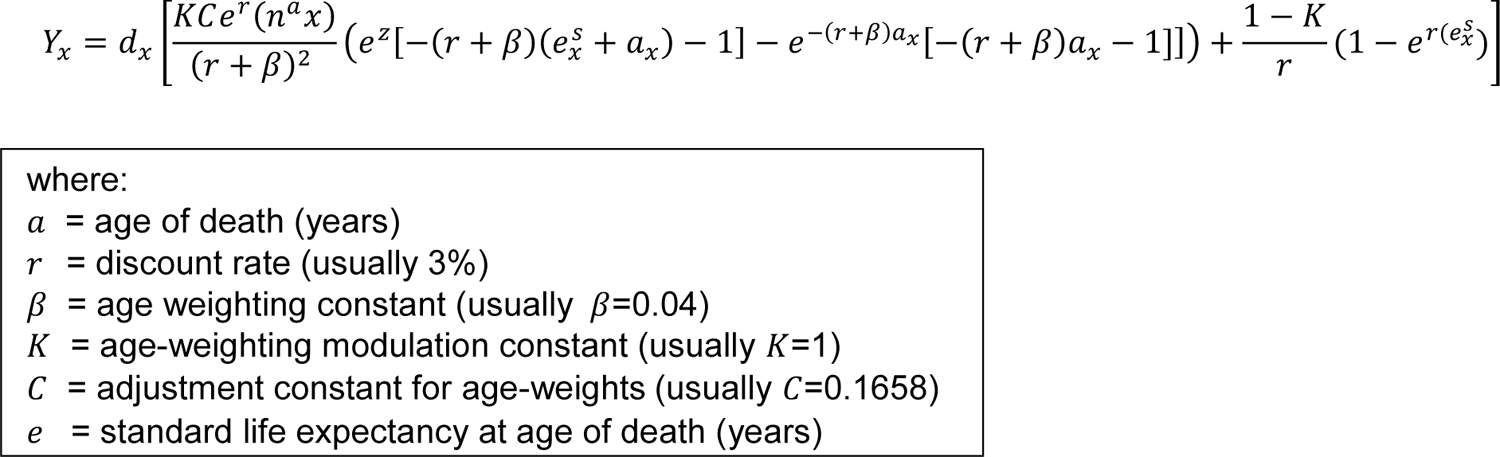

The discount rate (*r*) is based on the concept that future mortality outcomes have less value than the present mortality outcomes. The discounting rate is a continuous and exponential function, therefore the higher the discount rate, the lower the impact is in the future. The usual time for the discounting rate is set at 3% per year. Age weighting is used because it enables the YLL equation to value differently at various ages, i.e. the value of years lived by a young adult could be noted to be worth more than that of a very young or an old individual. So, the parameters were estimated at *C*=0.1658 and β=0.04 to account for this, which was calibrated and chosen from the 1990 GBD study.^1, 2^

There is an open source programming code available in R that includes this formula. Once the function is loaded, all that is required is a value of the number of death, average age of death, and the standard life expectancy for that age. The individual YLL is estimated. https://static-content.springer.com/esm/art%3A10.1186%2F1471-2458-8-116/MediaObjects/12889_2007_1086_MOESM3_ESM.pdf (Accessed 18-01-2022).

For an easier alternative to include all ages, an Excel calculation template can also be used from the World Health Organisation (WHO) National tools 2018, where the number of participants, deaths, and the standard life expectancy for that age can be entered into the template: https://www.who.int/quantifying_ehimpacts/publications/en/9241546204chap3.pdf (Accessed 18-01-2022).

The YLL for the UK Biobank example was calculated using the Excel template to get the full data. The YLL started at the age of 45 to 75+ in one year age intervals, for both men and women and with and without multimorbidity.

**Step 4.** The total YLL is summed, this can be aggregated.

**Step 5.** To find the average YLL, the total number of YLL is divided by the total number of deaths (in individual age groups if large enough, or in the aggregated age groups, i.e. 50-54 years).

**Step 6.** The average YLL for those with and without multimorbidity is subtracted to find the difference in YLL.

In our example, the average YLL was grouped into 5 year age group intervals to give an overall YLL, as the estimates for individual ages was too small. This method works best for large national data, as it is difficult to use when there are too few events and participants in the individual age groups.

## Methods S4. Chiang’s life tables example using UK Biobank

**Step 1.** Identify which life table to use.^1, 2^ The current life table was used for our example, as the UK Biobank included middle-aged participants, fewer deaths had occurred at younger ages than the older, and since the age structure is known to be affected by historical changes in fertility, mortality, and migration, which is unrelated to the current mortality.

Therefore, to avoid historical changes, a hypothetical cohort that is based on the observed probabilities of death at corresponding ages was used to reflect the effect on only mortality.

**Step 2.** Determine whether the life tables will be unabridged (i.e., a complete) life table or abridged life table. Unabridged life tables display data for every single year of age until the last age category, whist the abridge exhibit data by 5 or 10 year age intervals.

For our example, abridged life table with 5 year intervals was used starting from 45 years to the age of the oldest participant at follow-up, 81 years, since it has been recognised that large populations are needed to avoid systematic and random variations in mortality when constructing life tables.^2, 3^

**Step 3.** Determine the current age of participants as the age at last follow-up or the age at death and estimate the number of participants and deaths by age.

The number of deaths in each age interval was calculated for participants with and without multimorbidity.

**Step 4.** Construct the life table. The ONS provides further detailed information on the construction of National life tables.^1^

https://www.ons.gov.uk/ons/rel/subnational-health4/life-expec-at-birth-age-65/2004-06-to-2008-10/ref-life-table-template.xls (Accessed 20-10-2021).

The following gives a brief overview of the values for the life table:

*x* = the starting age of the interval.

*n* = width of the age interval.

*a*_*n*_ = defined to be 0.5, as we assume the deaths are evenly distributed over the age interval.

*m*_*x*_ = the rate of mortality, defined as the number of deaths at age *x (D_x_)* divided by the total population at that age (*P_x_*).

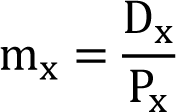

*q*_*x*_ = the mortality rate between age x and (x + 1), that is the conditional probability that a person aged x exact will die before reaching age (x + 1), given survival to age x.

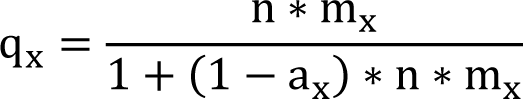

*p*_*x*_ = the survival rate between age x and (x + 1) based on the conditional probability.

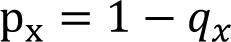

*l*_*x*_ = this is the number of survivors from a hypothetical cohort of new-born babies on which the life table is based. The initial value is arbitrary, i.e. 1, 100, 1000.

*d*_*x*_ = the number of deaths from a hypothetical cohort between exact age x and (x + 1).

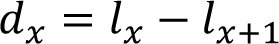

*L*_*x*_ = the number of person-years lived by a hypothetical cohort between exact age x and (*x* + 1).

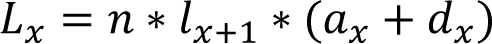

*T*_*x*_ = the number of person-years lived by a hypothetical cohort above exact age *x*, which is a cumulative distribution of the *L_x_* column.

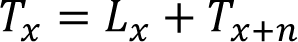

*e*_*x*_ = the expected number of years that those aged x exact will live thereafter in this hypothetical cohort.

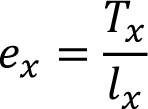

By calculating the variance of the conditional probability of death, the standard errors and the 95% confidence intervals (CI) can be estimated.

For these calculations, we used the ONS Excel template: https://www.ons.gov.uk/ons/rel/subnational-health4/life-expec-at-birth-age-65/2004-06-to-2008-10/ref-life-table-template.xls (Accessed 18-01-2022).

The number of participants and the number that have died by age groups and multimorbidity was inserted, and the hypothetical cohort started at 1000 (as shown in the Figure below).

**Step 5.** To find the difference in YLL between two groups, calculate two life tables separately (stratify) and then subtract the two expected number of years remaining.

The difference in YLL, which is age-group specific for the example, was calculated by subtracting the remaining life expectancy for those without multimorbidity and those with multimorbidity (for example, in the following Figure from the Excel template: YLL_45_ = 27.54 – 19.99 = 7.55; YLL_75_ = 3.50 – 3.50 = 0).

Excel template of example using Chiang’s life tables in those with and without multimorbidity:

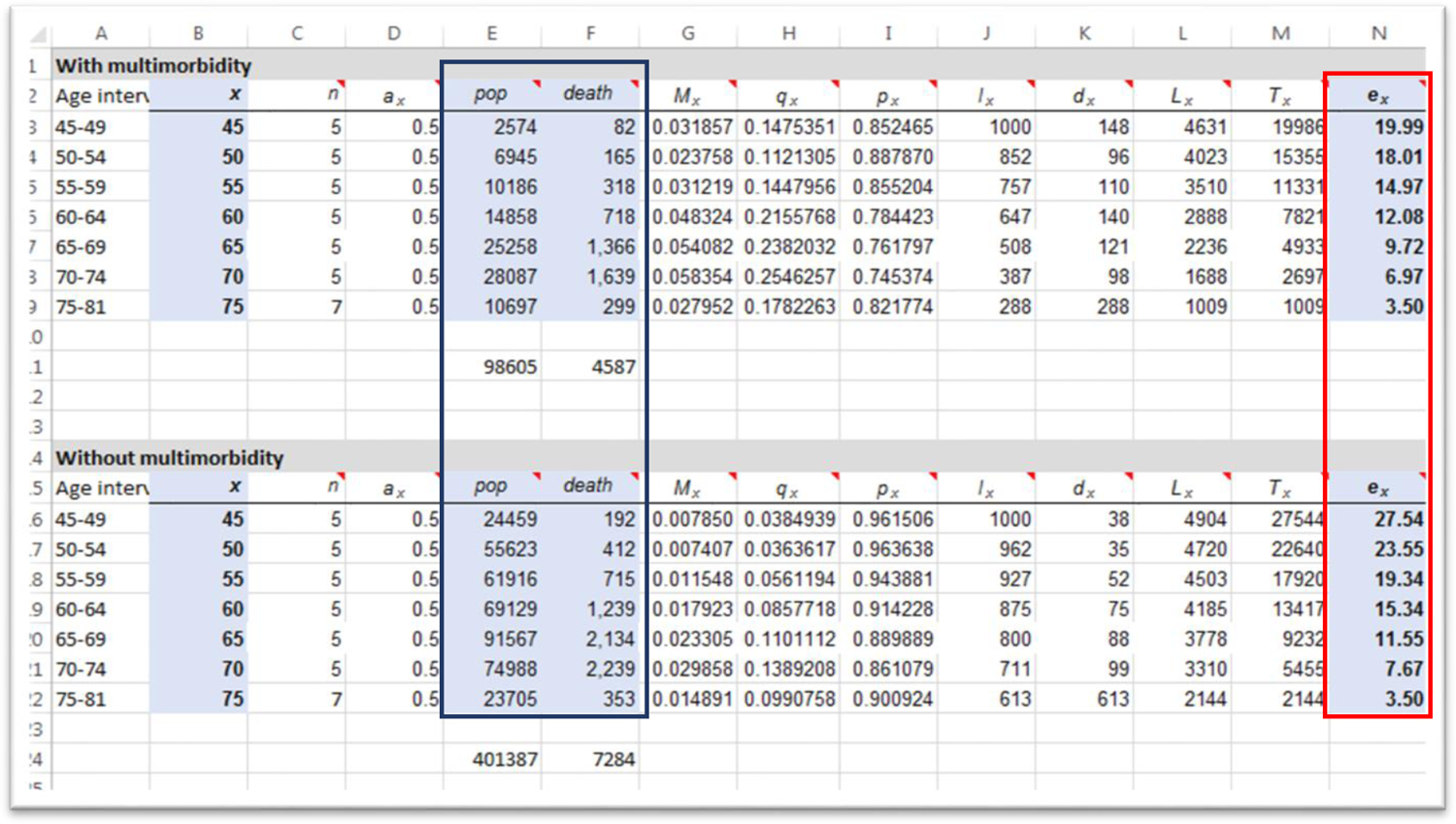

## Methods S5. Epi package example using UK Biobank

**Step 1.** Using the R software, install the Epi package and load the data.

**Step 2.** Estimate the survival curves for the two groups (multimorbidity and no multimorbidity) to quantify years of life lost (YLL) associated with having multimorbidity.

There are two ways to calculate the survival curves. In the Epi package, the example uses two groups in relation to the presence of diabetes: “well” (i.e., without diabetes) and “diseased” (i.e., with diabetes). It is possible to assume either that the “Well” persons are immune to diabetes, so persons without diabetes will not develop diabetes over time; or to model the transition rates between the “well” and “diseased” status, i.e., including the possibility that people without diabetes may develop diabetes over time. In our example, we will consider the first case (“immune”), as we want to compare subjects with multimorbidity to those without multimorbidity at baseline.

The details on the calculations are reported at 1) https://bendixcarstensen.com/ 2) http://bendixcarstensen.com/Epi/Courses/EDEG2017/ (Accessed 18-01-2022) and are briefly summarised here:

The survival function, for Well (W, no disease) and Disease (D) subjects, is derived from the instantaneous mortality rate (also known as the hazard function, hazard rate, *µ*) as follows:

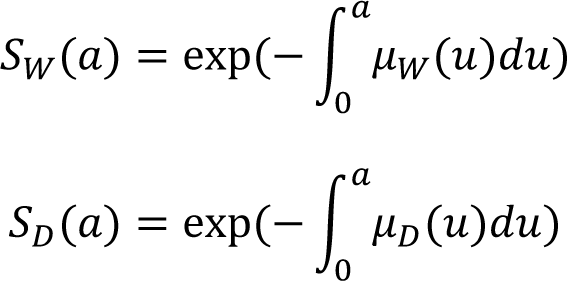

These are the survival functions from age 0 to age *a* in the two groups of subjects, with and without disease. We want conditional survival functions, given survival to age *A*:

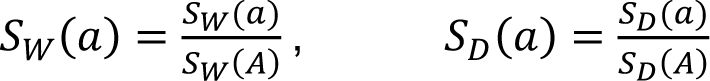

The age-specific mortality rates can be calculated using a Poisson model, after splitting the follow-up time in small intervals of age (“splitLexis” in the code below) and using a spline transformation of age as covariate. Rates are then predicted separately for subjects with and without multimorbidity, and subsequently used to obtain age-specific survival estimates (survival function) in each group.

The YLL is then calculated by integrating the survival functions with respect to age (*t*) of those well (no multimorbidity), *S*_*W*_(*t*), subtracted by those that have the disease (with multimorbidity), *S*_D_(*t*).

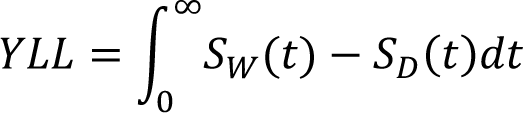

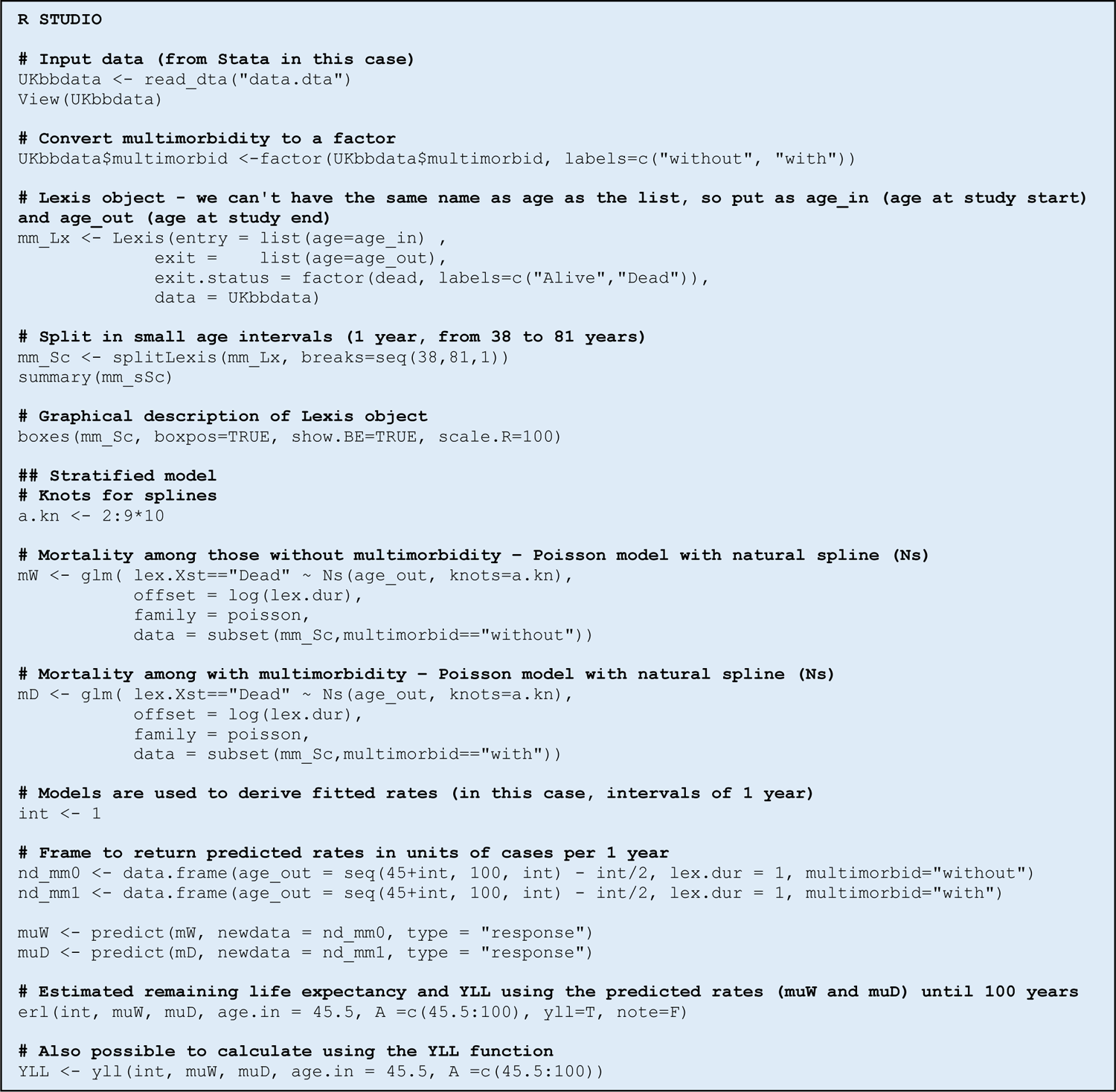

## Methods S6. Flexible Royston-Parmar parametric survival model example using UK Biobank

**Step 1.** Set up the data using *stset*: age is used as the time scale.

**Step 2.** Use the *stpm2* command to model survival data. Detailed explanations of the *stpm2* command can be found at: https://pclambert.net/software/stpm2/ (Accessed 18-01-2022).

Below is a brief explanation of the Royston-Parmar parametric survival model, implemented in the *stpm2* command.

A Cox proportional hazards model is defined as:

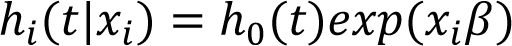

In the Cox model, we don’t estimate the baseline hazard ℎ_0_(*t*) – this cancels out in the partial likelihood – and we only estimate relative effects (i.e. hazard ratios). In the Royston-Parmar model, an additional parameters is required to estimate the baseline. In the case of the log cumulative hazard, the linear predictor is therefore:

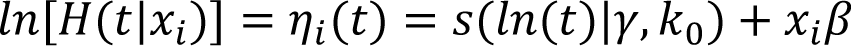

with *s(ln(*t*)|γ, k_0_)* being a restricted cubic spline function of log (time). Restricted cubic splines are used because they are forced to be linear before the first knot and after the final knot; this is where there is often less data and standard cubic splines tend to be sensitive to few extreme values.

**Step 3.** Use the *predictnl* command. This is used for postestimation predictions after fitting the model with *stpm2*. The command uses the delta method to calculate the variance (standard error, 95% confidence interval). The *surv* term (code below) indicates that the survival function is to be estimated for the group without (*at(multimorbid 0*)) and with (*at(multimorbid 1*)) multimorbidity. The *meansurv* is alternative to *surv* to obtain standardized (average) survival curves when including also covariates in the *stpm2* regression. Another command that does the two steps (*stmp2, meansurv*) together is *standsurv* (details are reported at: https://pclambert.net/software/standsurv/; Accessed 18-01-2022).

**Step 4.** Integrate the survival functions to find the remaining life expectancy for each group.

**Step 5.** YLL is estimated as the difference in the remaining life expectancy between the two groups.

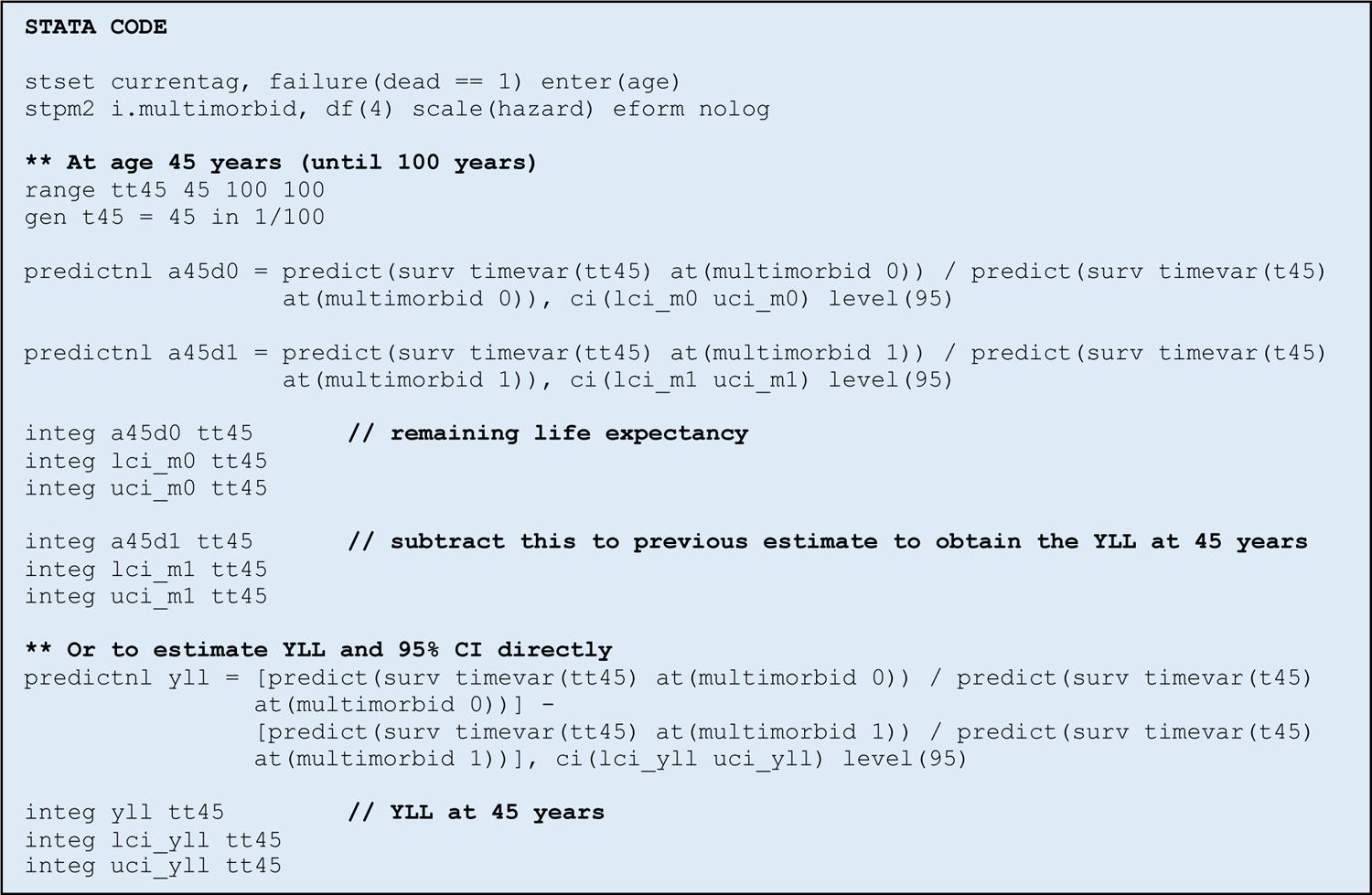

**Stata code to estimate years of life lost at different ages using flexible parametric Royston-Parmar survival model**

Multimorbidity can be considered as comparing any two groups to calculate the YLL.

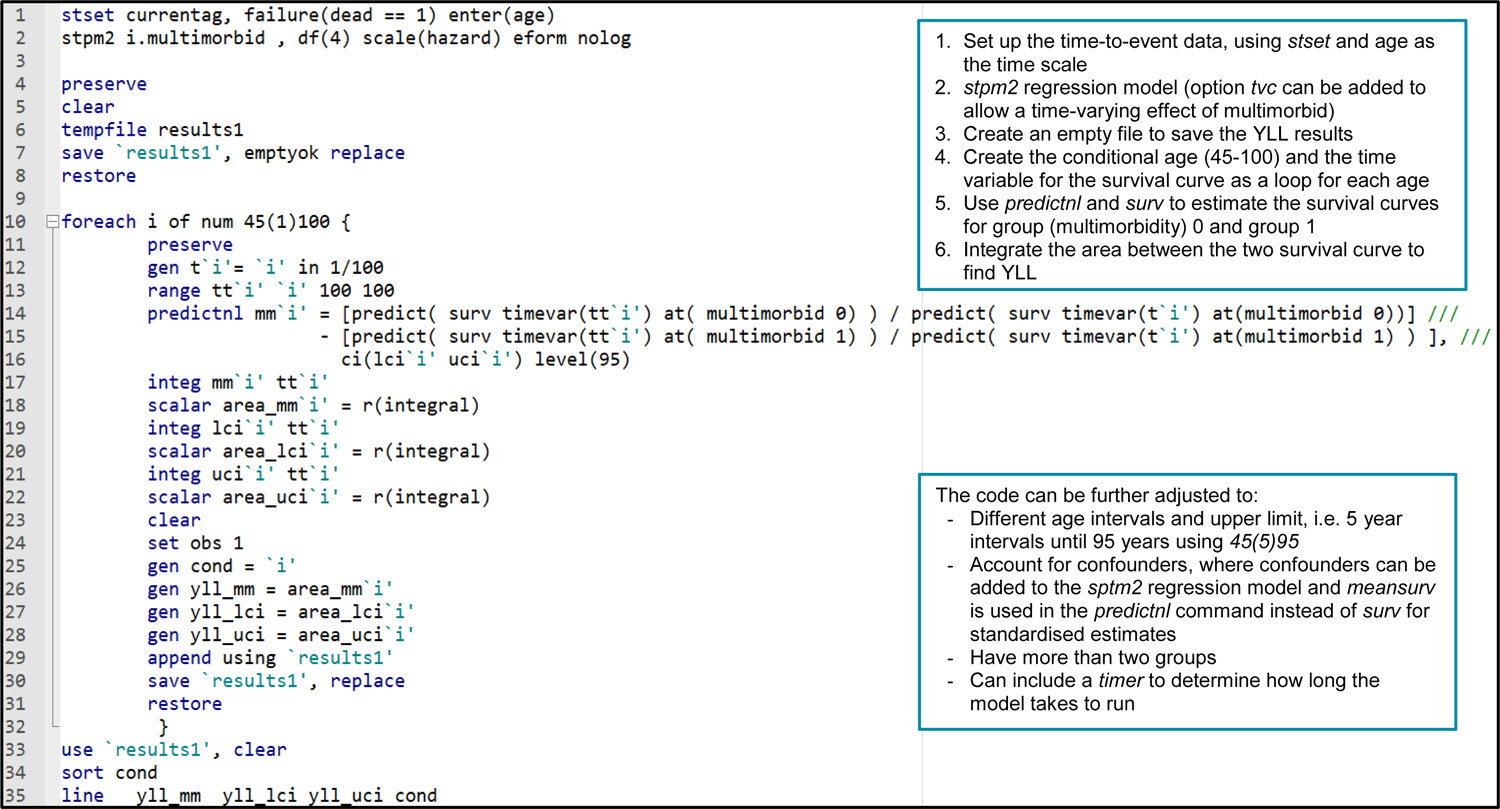

## Checklist S1. Strengthening the reporting of observational studies in epidemiology (STROBE)

|  | Item No | Recommendation | Manuscript |
| --- | --- | --- | --- |
| Title and abstract | 1 | (a) Indicate the study's design with a commonly used term in the title or the abstract | Title and Abstract |
|  |  | (b) Provide in the abstract an informative and balanced summary of what was done and what was found | Abstract |
| Introduction |  |  |  |
| Background/rationale | 2 | Explain the scientific background and rationale for the investigation being reported | Introduction paragraph 1-2 |
| Objectives | 3 | State specific objectives, including any prespecified hypotheses | Introduction paragraph 4 |
| Methods |  |  |  |
| Study design | 4 | Present key elements of study design early in the paper | Methods |
| Setting | 5 | Describe the setting, locations, and relevant dates, including periods of recruitment, exposure, follow-up, and data collection | Methods, Example from multimorbidity |
| Participants | 6 | (a) Cohort study—Give the eligibility criteria, and the sources and methods of selection of participants. Describe methods of follow-up | Methods, Example from multimorbidity |
|  |  | (b) Cohort study—For matched studies, give matching criteria and number of exposed and unexposed | NA |
| Variables | 7 | Clearly define all outcomes, exposures, predictors, potential confounders, and effect modifiers. Give diagnostic criteria, if applicable | Methods, Example from multimorbidity |
| Data sources/measurement | 8* | For each variable of interest, give sources of data and details of methods of assessment (measurement). Describe comparability of assessment methods if there is more than one group | Methods, Example from multimorbidity |
| Bias | 9 | Describe any efforts to address potential sources of bias | Methods, Example from multimorbidity |
| Study size | 10 | Explain how the study size was arrived at | Supplementary Figure S1 |
| Quantitative variables | 11 | Explain how quantitative variables were handled in the analyses. If applicable, describe which groupings were chosen and why | Methods, Example from multimorbidity |
| Statistical methods | 12 | (a) Describe all statistical methods, including those used to control for confounding | Methods, Example from multimorbidity |
|  |  | (b) Describe any methods used to examine subgroups and interactions | NA |
|  |  | (c) Explain how missing data were addressed | Methods, Example from multimorbidity |
|  |  | (d) Cohort study—If applicable, explain how loss to follow-up was addressed | NA |
|  |  | (e) Describe any sensitivity analyses | NA |
| Results |  |  |  |
| Participants | 13* | (a) Report numbers of individuals at each stage of study—e.g. numbers potentially eligible, examined for eligibility, confirmed eligible, included in the study, completing follow-up, and analysed | Methods, Example from multimorbidity |
|  |  | (b) Give reasons for non-participation at each stage | Methods, Example from multimorbidity |
|  |  | (c) Consider use of a flow diagram | Supplementary, Figure S1 |
| Descriptive data | 14* | (a) Give characteristics of study participants (e.g. demographic, clinical, social) and information on exposures and potential confounders | Results, Baseline Characteristics |
|  |  | (b) Indicate number of participants with missing data for each variable of interest | Supplementary, Figure S1 |
|  |  | (c) Cohort study—Summarise follow-up time (e.g., average and total amount) | NA |
| Outcome data | 15* | Cohort study—Report numbers of outcome events or summary measures over time | Results |
| Main results | 16 | (a) Give unadjusted estimates and, if applicable, confounder-adjusted estimates and their precision (eg, 95% confidence interval). Make clear which confounders were adjusted for and why they were included | Results |
|  |  | (b) Report category boundaries when continuous variables were categorized | Results |
|  |  | (c) If relevant, consider translating estimates of relative risk into absolute risk for a meaningful time period | Results |
| Other analyses | 17 | Report other analyses done—e.g. analyses of subgroups and interactions, and sensitivity analyses | NA |
| Discussion |  |  |  |
| Key results | 18 | Summarise key results with reference to study objectives | Discussion paragraph 1 |
| Limitations | 19 | Discuss limitations of the study, taking into account sources of potential bias or imprecision. Discuss both direction and magnitude of any potential bias | Discussion paragraph 8 |
| Interpretation | 20 | Give a cautious overall interpretation of results considering objectives, limitations, multiplicity of analyses, results from similar studies, and other relevant evidence | Discussion paragraph 2-9 |
| Generalisability | 21 | Discuss the generalisability (external validity) of the study results | Recommendations |
| Other information |  |  |  |
| Funding | 22 | Give the source of funding and the role of the funders for the present study and, if applicable, for the original study on which the present article is based. | End of the manuscript |

